# Identifying and Prioritizing Barriers to TB Prevention and Care in High-Burden Countries: A Community-Engaged Approach Using Best-Worst Scaling

**DOI:** 10.64898/2026.06.29.26356773

**Authors:** Andrew D. Kerkhoff, Kalee Singh, Nyiko Kubai, Mangala Namasivayam, Ketholelie Angami, Samyra R. Cox, Colleen F. Hanrahan, Thea Sigerman, Matthew Brandner, Erica Lessem, Priya Shete

## Abstract

**Objective:** To comprehensively identify the multi-level barriers to tuberculosis prevention and care in high-burden settings, and prioritize them according to their perceived harm and modifiability.

**Methods:** We conducted a multi-phase stakeholder-engaged study, synthesized barriers from two scoping reviews, and then convened regional workshops in Hyderabad, India, and Nairobi, Kenya, prioritizing TB survivors, community advocates, and frontline healthcare workers (HCWs). Participants refined existing barriers and added new ones. They then completed a Best-Worst Scaling (BWS) exercise to assess the perceived impact (harm) of each barrier, and Likert-based questions to assess perceived modifiability. Hierarchical Bayes (HB) modeling was used to generate mean impact scores (MIS) for each barrier, and mean scores for barrier modifiability (range, 1-4) were calculated.

**Findings:** 81 stakeholders from 28 countries participated, and 65 completed the survey (24% community representatives/advocates, 36% frontline HCWs; 17% were TB survivors). Stakeholder input expanded the barrier set from 31 to 39, with most newly added barriers at the health systems level. The BWS identified systems-level drug and supply challenges (MIS=5.8), patient/community-level financial factors (MIS=5.2), and inadequate provision of holistic care (MIS=4.9) as the highest-impact barriers; 7 of the top 10 barriers overlapped across regions. Asia participants placed 5 barriers, and Africa participants 10 in the high-impact/high-modifiability priority quadrant; four were shared across both: patient/community knowledge, HCW knowledge, HCW attitudes, and lack of community engagement mechanisms.

**Conclusion:** Incorporating the lived experience of TB-affected groups revealed substantial gaps in literature-derived TB care barriers. The high-impact, modifiable barriers identified provide actionable targets for programs in high-burden settings.

## Introduction

Despite being preventable and curable, tuberculosis (TB) is the leading infectious cause of death globally, killing more than 1.2 million people annually.^1^ This, in part, reflects persistent gaps between the availability of effective interventions and their successful delivery and uptake.^2–5^ The TB care cascade involves multiple touchpoints, from prevention among those at risk for TB, to symptom recognition, care-seeking, and treatment initiation and completion for people with TB.^6^ At each step, barriers operating at multiple levels - patient and community, healthcare worker (HCW), and health system - can undermine individual-level prevention and treatment outcomes as well as community- and country-level progress in the fight against TB.

Complex, multilevel barriers to TB prevention and care are well documented,^7–16^ with numerous studies from diverse contexts describing factors ranging from limited disease knowledge, TB-related stigma, and gendered factors at the community level, to inadequate knowledge and high workload among HCWs, to fragmented services and insufficient resources at the health system level. However, knowing which barriers exist is not sufficient to improve demand for, access to, and retention in TB services, and the sheer number of documented barriers can make prioritization difficult. Improving care requires both prioritizing which barriers to address and matching them to strategies suited to the local context.

Unfortunately, several knowledge gaps regarding key barriers to TB prevention and care hinder implementation progress in high-burden settings. First, although there is extensive literature on TB barriers, it is unknown how well it reflects the lived realities of people affected by TB and frontline HCWs rather than researcher framing. Second, many relevant studies are more than a decade old and may not accurately reflect the changes in post-COVID-19 service delivery or the contraction in global health funding in 2025 and its harmful effects on TB programs. Third, TB barriers have not been comprehensively synthesized and systematically prioritized based on the harm they cause or the feasibility of modifying them. Without this prioritization, programs often repeat the same activities, expecting different results, and fail to address key barriers, depleting precious resources without achieving meaningful progress.

To address these gaps, we conducted a stakeholder-engaged process with two major objectives: (1) to comprehensively identify barriers to TB prevention and care by integrating the perspectives of TB survivors, affected communities, and frontline HCWs with an evidence synthesis; and (2) to prioritize barriers by integrating their perceived harm with the feasibility of modifying them, identifying the most actionable barriers. These findings are intended to inform a global digital tool that matches priority barriers across diverse settings with the most promising implementation strategies to improve demand, access, and engagement in care across the TB care cascade.

## Methods

### Process overview

We conducted a multi-phase stakeholder-engaged process combining evidence synthesis with participatory refinement and preference elicitation. The process comprised two phases: (1) literature-based barrier identification from scoping reviews; and (2) regional stakeholder workshops for barrier refinement, expansion, and prioritization (**Box 1**).

### Phase 1: Initial barrier identification

We identified an initial set of barriers through two scoping reviews: an umbrella review synthesizing existing systematic reviews on barriers to TB prevention and care across the care cascade,^17^ and a focused review on barriers specific to HCWs involved in TB service delivery.^18^ The final set comprised 31 barriers: 12 at the patient/community level, 6 at the HCW level, and 13 at the health system level (**Table 1**). We characterized each barrier by: (1) its definition; (2) the step(s) of the TB care cascade it could directly affect (prevention, care-seeking, screening/case-finding, diagnosis, treatment initiation, and treatment adherence), recognizing that some plausibly affect several steps; (3) its influence on TB outcomes (reduced engagement/retention or poor clinical quality); and (4) the level it primarily acts on: patient/community, HCW, or health system. We use “patient/community level” as a framework label encompassing people affected by TB, those at risk, and the wider community. Because HCW behaviors are substantially shaped by their working environment, we defaulted to a system-level classification when attribution was ambiguous.

**Table 1.**
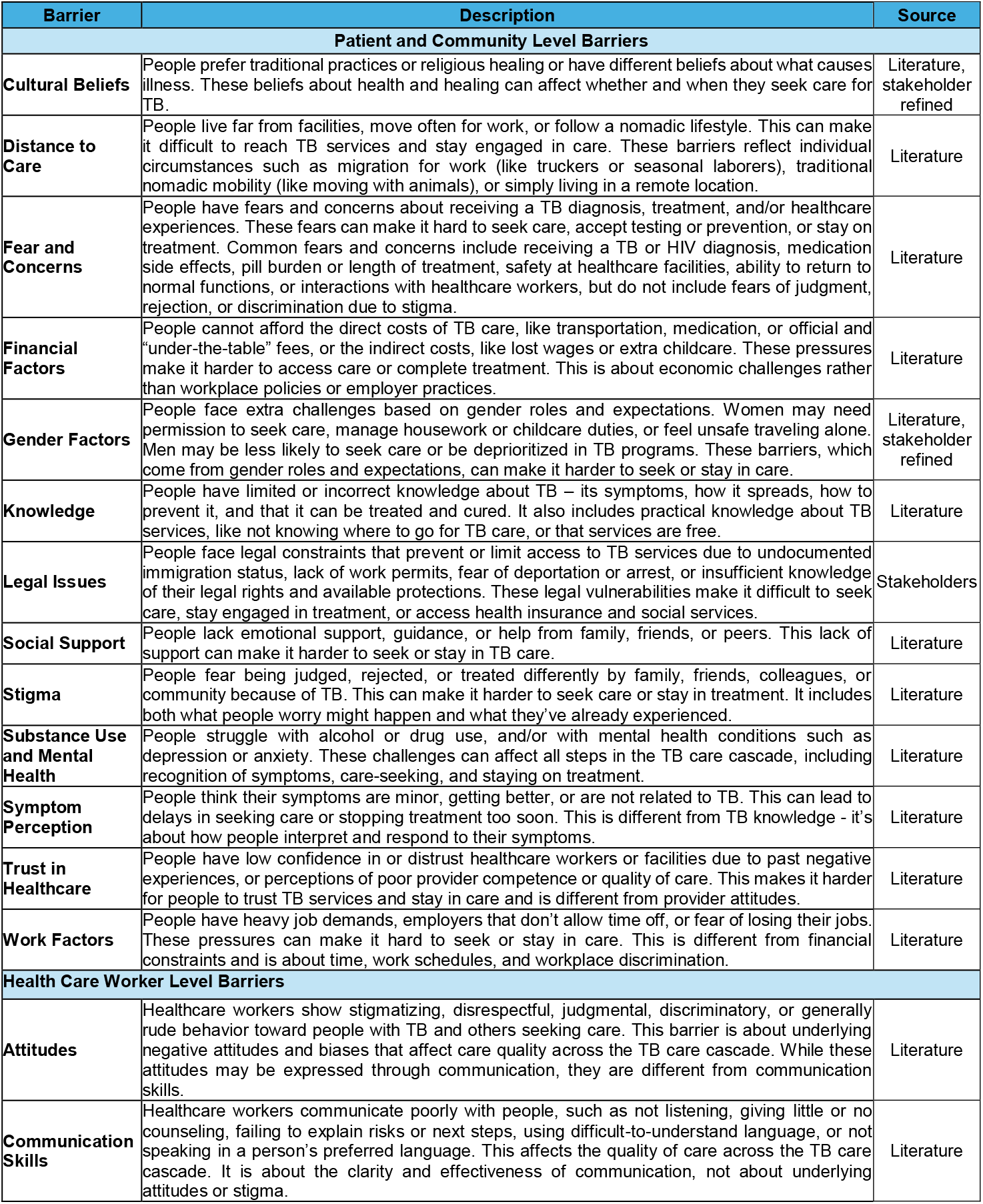
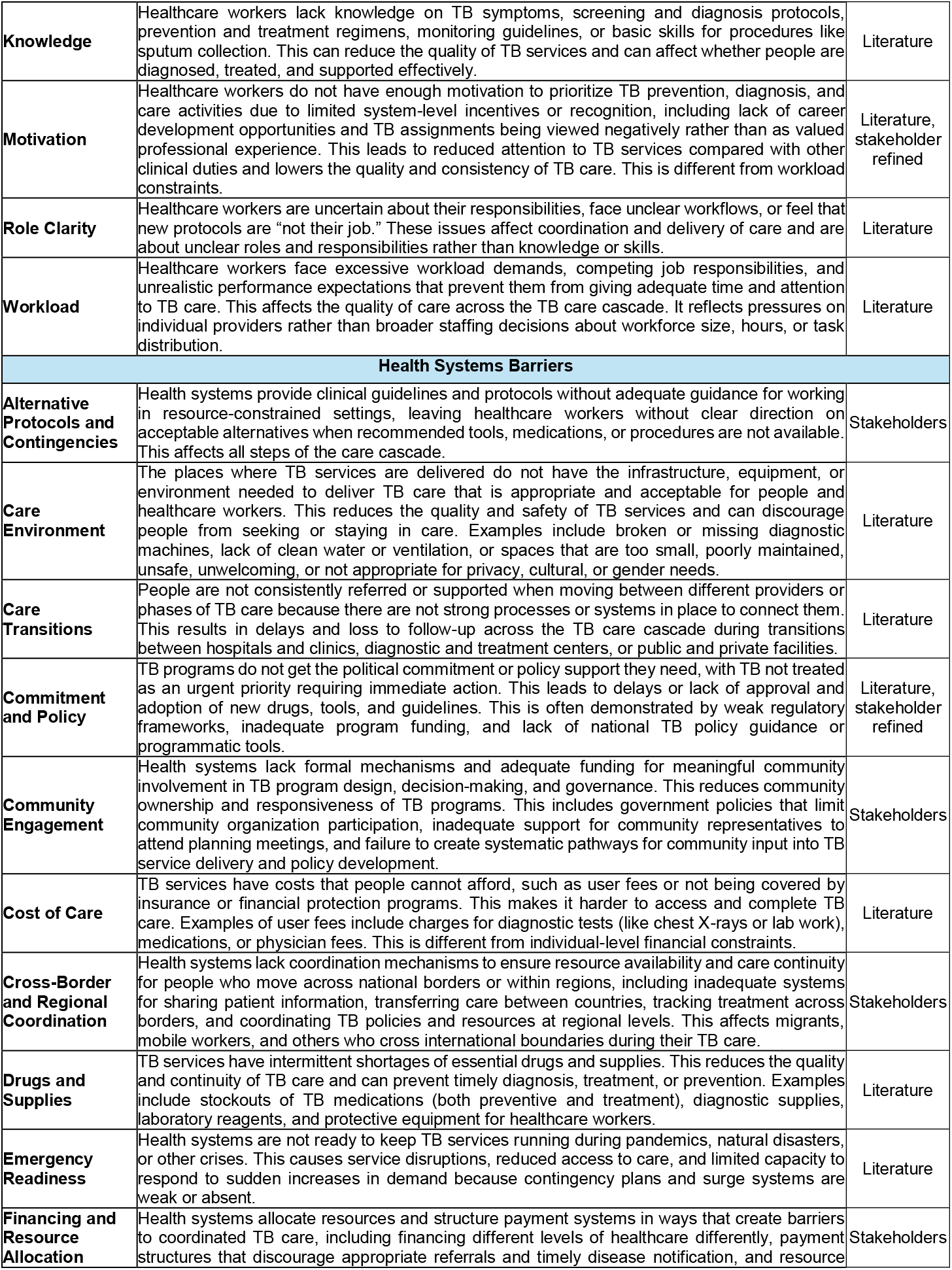
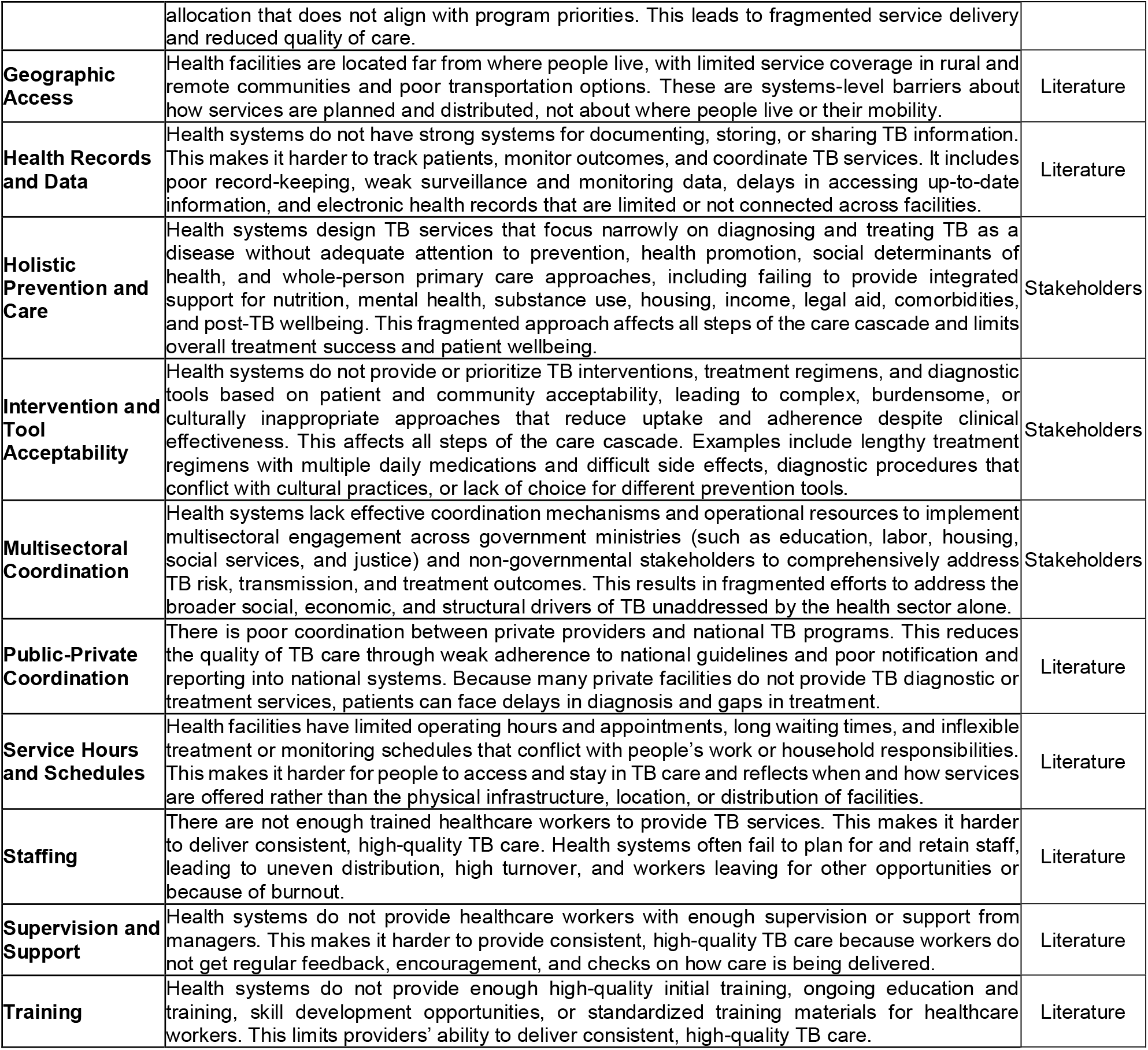
Barriers to TB prevention and care informed by the literature and stakeholders. Barriers are categorized as literature-derived, literature-derived with stakeholder refinements (either scope or description), or stakeholder-derived.

| Barrier | Description | Source |
| --- | --- | --- |
| **Patient and Community Level Barriers** |  |  |
| **Cultural Beliefs** | People prefer traditional practices or religious healing or have different beliefs about what causes illness. These beliefs about health and healing can affect whether and when they seek care for TB. | Literature, stakeholder refined |
| **Distance to Care** | People live far from facilities, move often for work, or follow a nomadic lifestyle. This can make it difficult to reach TB services and stay engaged in care. These barriers reflect individual circumstances such as migration for work (like truckers or seasonal laborers), traditional nomadic mobility (like moving with animals), or simply living in a remote location. | Literature |
| **Fear and Concerns** | People have fears and concerns about receiving a TB diagnosis, treatment, and/or healthcare experiences. These fears can make it hard to seek care, accept testing or prevention, or stay on treatment. Common fears and concerns include receiving a TB or HIV diagnosis, medication side effects, pill burden or length of treatment, safety at healthcare facilities, ability to return to normal functions, or interactions with healthcare workers, but do not include fears of judgment, rejection, or discrimination due to stigma. | Literature |
| **Financial Factors** | People cannot afford the direct costs of TB care, like transportation, medication, or official and “under-the-table” fees, or the indirect costs, like lost wages or extra childcare. These pressures make it harder to access care or complete treatment. This is about economic challenges rather than workplace policies or employer practices. | Literature |
| **Gender Factors** | People face extra challenges based on gender roles and expectations. Women may need permission to seek care, manage housework or childcare duties, or feel unsafe traveling alone. Men may be less likely to seek care or be deprioritized in TB programs. These barriers, which come from gender roles and expectations, can make it harder to seek or stay in care. | Literature, stakeholder refined |
| **Knowledge** | People have limited or incorrect knowledge about TB – its symptoms, how it spreads, how to prevent it, and that it can be treated and cured. It also includes practical knowledge about TB services, like not knowing where to go for TB care, or that services are free. | Literature |
| **Legal Issues** | People face legal constraints that prevent or limit access to TB services due to undocumented immigration status, lack of work permits, fear of deportation or arrest, or insufficient knowledge of their legal rights and available protections. These legal vulnerabilities make it difficult to seek care, stay engaged in treatment, or access health insurance and social services. | Stakeholders |
| **Social Support** | People lack emotional support, guidance, or help from family, friends, or peers. This lack of support can make it harder to seek or stay in TB care. | Literature |
| **Stigma** | People fear being judged, rejected, or treated differently by family, friends, colleagues, or community because of TB. This can make it harder to seek care or stay in treatment. It includes both what people worry might happen and what they’ve already experienced. | Literature |
| **Substance Use and Mental Health** | People struggle with alcohol or drug use, and/or with mental health conditions such as depression or anxiety. These challenges can affect all steps in the TB care cascade, including recognition of symptoms, care-seeking, and staying on treatment. | Literature |
| **Symptom Perception** | People think their symptoms are minor, getting better, or are not related to TB. This can lead to delays in seeking care or stopping treatment too soon. This is different from TB knowledge - it’s about how people interpret and respond to their symptoms. | Literature |
| **Trust in Healthcare** | People have low confidence in or distrust healthcare workers or facilities due to past negative experiences, or perceptions of poor provider competence or quality of care. This makes it harder for people to trust TB services and stay in care and is different from provider attitudes. | Literature |
| **Work Factors** | People have heavy job demands, employers that don’t allow time off, or fear of losing their jobs. These pressures can make it hard to seek or stay in care. This is different from financial constraints and is about time, work schedules, and workplace discrimination. | Literature |
| **Health Care Worker Level Barriers** |  |  |
| **Attitudes** | Healthcare workers show stigmatizing, disrespectful, judgmental, discriminatory, or generally rude behavior toward people with TB and others seeking care. This barrier is about underlying negative attitudes and biases that affect care quality across the TB care cascade. While these attitudes may be expressed through communication, they are different from communication skills. | Literature |
| **Communication Skills** | Healthcare workers communicate poorly with people, such as not listening, giving little or no counseling, failing to explain risks or next steps, using difficult-to-understand language, or not speaking in a person’s preferred language. This affects the quality of care across the TB care cascade. It is about the clarity and effectiveness of communication, not about underlying attitudes or stigma. | Literature |
| **Knowledge** | Healthcare workers lack knowledge on TB symptoms, screening and diagnosis protocols, prevention and treatment regimens, monitoring guidelines, or basic skills for procedures like sputum collection. This can reduce the quality of TB services and can affect whether people are diagnosed, treated, and supported effectively. | Literature |
| **Motivation** | Healthcare workers do not have enough motivation to prioritize TB prevention, diagnosis, and care activities due to limited system-level incentives or recognition, including lack of career development opportunities and TB assignments being viewed negatively rather than as valued professional experience. This leads to reduced attention to TB services compared with other clinical duties and lowers the quality and consistency of TB care. This is different from workload constraints. | Literature, stakeholder refined |
| **Role Clarity** | Healthcare workers are uncertain about their responsibilities, face unclear workflows, or feel that new protocols are “not their job.” These issues affect coordination and delivery of care and are about unclear roles and responsibilities rather than knowledge or skills. | Literature |
| **Workload** | Healthcare workers face excessive workload demands, competing job responsibilities, and unrealistic performance expectations that prevent them from giving adequate time and attention to TB care. This affects the quality of care across the TB care cascade. It reflects pressures on individual providers rather than broader staffing decisions about workforce size, hours, or task distribution. | Literature |
| **Health Systems Barriers** |  |  |
| **Alternative Protocols and Contingencies** | Health systems provide clinical guidelines and protocols without adequate guidance for working in resource-constrained settings, leaving healthcare workers without clear direction on acceptable alternatives when recommended tools, medications, or procedures are not available. This affects all steps of the care cascade. | Stakeholders |
| **Care Environment** | The places where TB services are delivered do not have the infrastructure, equipment, or environment needed to deliver TB care that is appropriate and acceptable for people and healthcare workers. This reduces the quality and safety of TB services and can discourage people from seeking or staying in care. Examples include broken or missing diagnostic machines, lack of clean water or ventilation, or spaces that are too small, poorly maintained, unsafe, unwelcoming, or not appropriate for privacy, cultural, or gender needs. | Literature |
| **Care Transitions** | People are not consistently referred or supported when moving between different providers or phases of TB care because there are not strong processes or systems in place to connect them. This results in delays and loss to follow-up across the TB care cascade during transitions between hospitals and clinics, diagnostic and treatment centers, or public and private facilities. | Literature |
| **Commitment and Policy** | TB programs do not get the political commitment or policy support they need, with TB not treated as an urgent priority requiring immediate action. This leads to delays or lack of approval and adoption of new drugs, tools, and guidelines. This is often demonstrated by weak regulatory frameworks, inadequate program funding, and lack of national TB policy guidance or programmatic tools. | Literature, stakeholder refined |
| **Community Engagement** | Health systems lack formal mechanisms and adequate funding for meaningful community involvement in TB program design, decision-making, and governance. This reduces community ownership and responsiveness of TB programs. This includes government policies that limit community organization participation, inadequate support for community representatives to attend planning meetings, and failure to create systematic pathways for community input into TB service delivery and policy development. | Stakeholders |
| **Cost of Care** | TB services have costs that people cannot afford, such as user fees or not being covered by insurance or financial protection programs. This makes it harder to access and complete TB care. Examples of user fees include charges for diagnostic tests (like chest X-rays or lab work), medications, or physician fees. This is different from individual-level financial constraints. | Literature |
| **Cross-Border and Regional Coordination** | Health systems lack coordination mechanisms to ensure resource availability and care continuity for people who move across national borders or within regions, including inadequate systems for sharing patient information, transferring care between countries, tracking treatment across borders, and coordinating TB policies and resources at regional levels. This affects migrants, mobile workers, and others who cross international boundaries during their TB care. | Stakeholders |
| **Drugs and Supplies** | TB services have intermittent shortages of essential drugs and supplies. This reduces the quality and continuity of TB care and can prevent timely diagnosis, treatment, or prevention. Examples include stockouts of TB medications (both preventive and treatment), diagnostic supplies, laboratory reagents, and protective equipment for healthcare workers. | Literature |
| **Emergency Readiness** | Health systems are not ready to keep TB services running during pandemics, natural disasters, or other crises. This causes service disruptions, reduced access to care, and limited capacity to respond to sudden increases in demand because contingency plans and surge systems are weak or absent. | Literature |
| **Financing and Resource Allocation** | Health systems allocate resources and structure payment systems in ways that create barriers to coordinated TB care, including financing different levels of healthcare differently, payment structures that discourage appropriate referrals and timely disease notification, and resource | Stakeholders |
|  | allocation that does not align with program priorities. This leads to fragmented service delivery and reduced quality of care. |  |
| **Geographic Access** | Health facilities are located far from where people live, with limited service coverage in rural and remote communities and poor transportation options. These are systems-level barriers about how services are planned and distributed, not about where people live or their mobility. | Literature |
| **Health Records and Data** | Health systems do not have strong systems for documenting, storing, or sharing TB information. This makes it harder to track patients, monitor outcomes, and coordinate TB services. It includes poor record-keeping, weak surveillance and monitoring data, delays in accessing up-to-date information, and electronic health records that are limited or not connected across facilities. | Literature |
| **Holistic Prevention and Care** | Health systems design TB services that focus narrowly on diagnosing and treating TB as a disease without adequate attention to prevention, health promotion, social determinants of health, and whole-person primary care approaches, including failing to provide integrated support for nutrition, mental health, substance use, housing, income, legal aid, comorbidities, and post-TB wellbeing. This fragmented approach affects all steps of the care cascade and limits overall treatment success and patient wellbeing. | Stakeholders |
| **Intervention and Tool Acceptability** | Health systems do not provide or prioritize TB interventions, treatment regimens, and diagnostic tools based on patient and community acceptability, leading to complex, burdensome, or culturally inappropriate approaches that reduce uptake and adherence despite clinical effectiveness. This affects all steps of the care cascade. Examples include lengthy treatment regimens with multiple daily medications and difficult side effects, diagnostic procedures that conflict with cultural practices, or lack of choice for different prevention tools. | Stakeholders |
| **Multisectoral Coordination** | Health systems lack effective coordination mechanisms and operational resources to implement multisectoral engagement across government ministries (such as education, labor, housing, social services, and justice) and non-governmental stakeholders to comprehensively address TB risk, transmission, and treatment outcomes. This results in fragmented efforts to address the broader social, economic, and structural drivers of TB unaddressed by the health sector alone. | Stakeholders |
| **Public-Private Coordination** | There is poor coordination between private providers and national TB programs. This reduces the quality of TB care through weak adherence to national guidelines and poor notification and reporting into national systems. Because many private facilities do not provide TB diagnostic or treatment services, patients can face delays in diagnosis and gaps in treatment. | Literature |
| **Service Hours and Schedules** | Health facilities have limited operating hours and appointments, long waiting times, and inflexible treatment or monitoring schedules that conflict with people's work or household responsibilities. This makes it harder for people to access and stay in TB care and reflects when and how services are offered rather than the physical infrastructure, location, or distribution of facilities. | Literature |
| **Staffing** | There are not enough trained healthcare workers to provide TB services. This makes it harder to deliver consistent, high-quality TB care. Health systems often fail to plan for and retain staff, leading to uneven distribution, high turnover, and workers leaving for other opportunities or because of burnout. | Literature |
| **Supervision and Support** | Health systems do not provide healthcare workers with enough supervision or support from managers. This makes it harder to provide consistent, high-quality TB care because workers do not get regular feedback, encouragement, and checks on how care is being delivered. | Literature |
| **Training** | Health systems do not provide enough high-quality initial training, ongoing education and training, skill development opportunities, or standardized training materials for healthcare workers. This limits providers' ability to deliver consistent, high-quality TB care. | Literature |

### Phase 2: Regional Stakeholder Workshops

#### Overview of workshops and participants

We convened two regional workshops, an Asia workshop in Hyderabad, India (September 22-23, 2025), and an Africa workshop in Nairobi, Kenya (October 14-15, 2025), each spanning two days. Day 1 focused on refining, expanding, and prioritizing barriers, while Day 2 focused on evaluating candidate implementation strategies to address the prioritized barriers (reported elsewhere). We deliberately prioritized participation from groups typically underrepresented in TB policy discussions: TB survivors, community advocates, and frontline HCWs. Participants discussed, refined, and expanded the barrier set based on their experience and expertise, and then completed a Best-Worst Scaling (BWS) exercise and modifiability assessment of the barriers. The workshops are described in detail in **Box 1**.

### Best-Worst Scaling and Survey Design

The impact and modifiability assessment were embedded in a self-completed online survey covering up to 39 barriers; two were identified during the later Africa workshop and assessed only by African participants, while 37 were assessed in both workshops. We selected BWS (type 1) over alternatives (e.g., rankings or ratings) to assess the relative perceived impact of barriers, because it provides more robust discrimination among a large number of items while reducing cognitive burden.^19,20^ The BWS exercise was designed using Sawtooth Software (Discover) and had a near-balanced incomplete block design.^21^ Participants were shown 28 and 30 choice tasks in the Asia and Africa workshops, respectively, to ensure each barrier was seen three times, allowing for stable individual-level estimates. In each choice task, participants viewed four randomly drawn barriers and selected the one they perceived as causing the “most harm” to good or high-quality TB outcomes in their setting and the one causing the “least harm,” independent of perceived modifiability (**Supplementary Figure 1a**).

Participants also rated the perceived modifiability of each barrier on a 4-point Likert scale ranging from 1 (not modifiable) to 4 (easy to modify) (**Supplementary Figure 1b**), assessing modifiability independently of perceived harm. They were asked to consider barriers in the context of settings they know and work in, reflecting on typically available resources, staff, and funding; required approvals or partnerships; existing strengths that could be built upon; and whether changes could realistically be carried out and sustained.

### Statistical analysis

BWS data were analyzed using a Hierarchical Bayes (HB) model in Sawtooth Software (Discover),^22^ to estimate individual-level preference weights for each barrier. These were rescaled to sum to 100 across all barriers to improve interpretation; we subsequently refer to these as “mean impact scores (MIS).” Mean scores (1-4) were also calculated for modifiability ratings. Estimates were determined overall and by region. To generate pooled overall estimates, BWS results for African participants were rescaled to the 37 common barriers assessed in both workshops. Regional estimates used the full barrier set for each workshop (37 for Asia and 39 for Africa), enabling Africa-specific barriers to be positioned within each region’s priority matrix while still allowing cross-regional comparisons. We combined BWS-derived impact scores with Likert-derived modifiability ratings to classify barriers into a 2×2 priority matrix for each region. A barrier was classified as a priority based on impact if its MIS exceeded the overall mean (indicating greater-than-average perceived harm) and as a priority based on modifiability if its mean Likert score was 3 or higher (indicating the barrier was perceived as at least somewhat modifiable).

### Ethics statement

The University of California, San Francisco Institutional Review Board determined this study did not meet the threshold for human subjects research (#26-45819).

## Results

### Workshop participant characteristics

Overall, 81 stakeholders from 28 countries, including 23 designated by WHO as high-burden for TB, HIV-associated TB, or MDR/RR-TB, participated across the two workshops (43 in Asia, 38 in Africa; 47 [58%] in person; **Supplementary Table 1**). Of the 65 who completed the survey-based barrier assessment (32 in Asia, 33 in Africa), 25% were community representatives, 35% frontline HCWs, 20% policy/decision makers, and 15% researchers; 17% reported being a TB survivor, 57% reported 10+ years of paid or unpaid work in the TB field, 43% reported involvement in national TB program or policy decision-making, and 36% worked primarily in urban settings, 6% in rural settings, and 58% in both (**Supplementary Table 2**).

### Barrier identification outcomes

Stakeholder input during the workshops expanded the barrier set from 31 literature-derived barriers to 39 total barriers (8 new barriers; 21% of total). The Asia workshop identified 6 new barriers (1 at the patient/community level and 5 at the health system level), while the Africa workshop identified 2 additional barriers (both at the health system level). The 39 final barriers were distributed across levels as follows: 13 at the patient/community level, 6 at the HCW level, and 20 at the health system level. These are summarized, along with their definitions refined through stakeholder feedback, in **Table 1**.

### Barrier impact, modifiability, and prioritization

Across the 37 common barriers, MIS ranged from 0.8 to 5.8 (mean 2.7, standard deviation 1.3). Those perceived as causing the greatest harm to TB outcomes were drugs and supplies (systems-level; MIS=5.8), financial factors (patient/community-level; MIS=5.2), lack of holistic prevention and care (systems-level; MIS=4.9), financing and resource allocation (systems-level; MIS=4.7), and lack of community engagement mechanisms (systems-level; MIS=4.7; **Table 2**). Health system-level barriers comprised 6 of the top 10 highest-impact barriers. Despite small differences in the barrier sets evaluated (37 common barriers, plus 2 Africa-specific barriers), there was substantial concordance in prioritization across regions, with seven of the top 10 highest-impact barriers being consistent across Asia and Africa workshops **(Supplementary Table 2)**. Of the two additional barriers evaluated only during the Africa workshop, multisectoral coordination and cross-border and regional coordination ranked 8^th^ and 16^th^ (of 39), respectively.

**Table 2.** Overall TB barrier impact and modifiability scores pooled across both regions (n=65). Mean impact scores (MIS) represent BWS-derived mean preference weights and sum to 100 across all barriers. A higher MIS indicates greater perceived harm; because all barriers are on the same scale, MIS can be directly compared (e.g., barrier A is twice as harmful as barrier B). Mean modifiability scores range from 1 (“not modifiable”) to 4 (easy to modify), with score of 3 indicating a barrier was, on average, perceived as somewhat modifiable.

| Barrier | Level | Overall (n=65) |  |  |
| --- | --- | --- | --- | --- |
|  |  | Impact Rank | Mean Impact Score | Mean Modifiability Score |
| Drugs & supplies | Systems | 1 | 5.8 (5.2-6.3) | 3.0 (2.8-3.2) |
| Financial factors | Patient | 2 | 5.2 (4.7-5.7) | 2.6 (2.4-2.8) |
| Holistic prevention & care | Systems | 3 | 4.9 (4.3-5.5) | 2.9 (2.7-3.1) |
| Financing & resource allocation | Systems | 4 | 4.7 (4.2-5.2) | 2.5 (2.4-2.7) |
| Community engagement | Systems | 5 | 4.7 (4.1-5.3) | 3.2 (3.0-3.4) |
| Stigma | Patient | 6 | 4.2 (3.6-4.8) | 2.9 (2.8-3.1) |
| Commitment & policy | Systems | 7 | 4.0 (3.3-4.6) | 2.6 (2.4-2.8) |
| Cost of care | Systems | 8 | 3.8 (3.2-4.4) | 2.7 (2.5-2.9) |
| Knowledge | Patient | 9 | 3.7 (3.2-4.2) | 3.7 (3.6-3.8) |
| Knowledge | HCW | 10 | 3.5 (3.0-4.0) | 3.7 (3.6-3.8) |
| Attitudes | HCW | 11 | 3.4 (2.8-3.9) | 3.2 (3.0-3.3) |
| Intervention acceptability | Systems | 12 | 3.4 (2.9-3.9) | 3.0 (2.8-3.2) |
| Symptom perception | Patient | 13 | 2.9 (2.3-3.5) | 3.4 (3.3-3.6) |
| Care environment | Systems | 14 | 2.9 (2.4-3.4) | 2.9 (2.7-3.0) |
| Trust in healthcare | Patient | 15 | 2.8 (2.3-3.3) | 3.2 (3.1-3.4) |
| Communication skills | HCW | 16 | 2.8 (2.2-3.3) | 3.5 (3.4-3.7) |
| Workload | HCW | 17 | 2.5 (2.1-3.0) | 2.8 (2.6-2.9) |
| Substance use & mental health | Patient | 18 | 2.5 (2.0-3.0) | 2.6 (2.4-2.8) |
| Geographic access | Systems | 19 | 2.5 (2.0-3.0) | 2.6 (2.4-2.8) |
| Staffing | Systems | 20 | 2.3 (1.8-2.8) | 3.1 (2.9-3.2) |
| Fear & concerns | Patient | 21 | 2.2 (1.7-2.6) | 3.4 (3.2-3.5) |
| Motivation | HCW | 22 | 2.0 (1.6-2.5) | 3.0 (2.8-3.2) |
| Social support | Patient | 23 | 2.0 (1.5-2.5) | 3.2 (3.0-3.3) |
| Distance to care | Patient | 24 | 1.9 (1.5-2.3) | 2.7 (2.5-2.9) |
| Emergency readiness | Systems | 25 | 1.9 (1.4-2.4) | 2.8 (2.6-3.0) |
| Alternative protocols | Systems | 26 | 1.9 (1.5-2.2) | 3.1 (3.0-3.3) |
| Training | Systems | 27 | 1.8 (1.5-2.1) | 3.4 (3.3-3.6) |
| Service hours & schedules | Systems | 28 | 1.8 (1.4-2.2) | 3.1 (2.9-3.3) |
| Cultural beliefs | Patient | 29 | 1.7 (1.3-2.1) | 2.7 (2.5-2.9) |
| Public-private coordination | Systems | 30 | 1.6 (1.2-2.1) | 3.0 (2.8-3.2) |
| Care transitions | Systems | 31 | 1.6 (1.1-2.0) | 3.2 (3.0-3.4) |
| Legal issues | Patient | 32 | 1.5 (1.0-2.1) | 2.6 (2.3-2.8) |
| Work factors | Patient | 33 | 1.5 (1.1-1.9) | 2.7 (2.5-2.9) |
| Gender factors | Patient | 34 | 1.5 (1.1-1.9) | 3.0 (2.9-3.2) |
| Supervision & support | Systems | 35 | 1.1 (0.7-1.5) | 3.3 (3.2-3.5) |
| Health records & data | Systems | 36 | 0.9 (0.7-1.2) | 3.2 (3.0-3.4) |
| Role clarity | HCW | 37 | 0.8 (0.6-1.1) | 3.5 (3.4-3.7) |

Note that cross-border & regional coordination and multisectoral coordination are not shown as these were not evaluated by participants in the Asia workshop.

Overall, 21 (57%) of the 37 common barriers had a mean modifiability score of 3 or higher (indicating they were perceived as at least somewhat modifiable), while no barrier had a mean score of 2 or lower (indicating perception as very difficult or not modifiable) (**Table 2)**. There was substantial concordance in barrier modifiability ratings across regions **(Supplementary Table 2**).

Priority matrices combining the perceived impact (i.e., harm) and modifiability of each barrier are shown by region in **Figure 1**. Five barriers in Asia and 10 in Africa fell into the high-impact/high-modifiability quadrant, representing immediate priorities for implementation efforts. Four barriers appeared in the priority quadrant across both regions: patient/community knowledge, HCW knowledge, HCW attitudes, and systems-level lack of community engagement mechanisms. Beyond these common priorities, Asia’s immediate priority quadrant included trust in healthcare (patient/community-level), while Africa’s included stigma (patient/community-level), symptom perception (patient/community-level), HCW communication skills, holistic prevention & care (systems-level), intervention acceptability (systems-level), and drugs & supplies (systems-level). Although drugs & supplies was one of the two highest-impact barriers in both regions, it fell just outside the immediate priority quadrant in Asia due to slightly lower perceived modifiability. Among the 10 highest-impact barriers in each region, 7 in Asia fell into the strategic opportunities quadrant (including the top 3), compared to 4 in Africa (none of the top 3).

**Figure 1.**
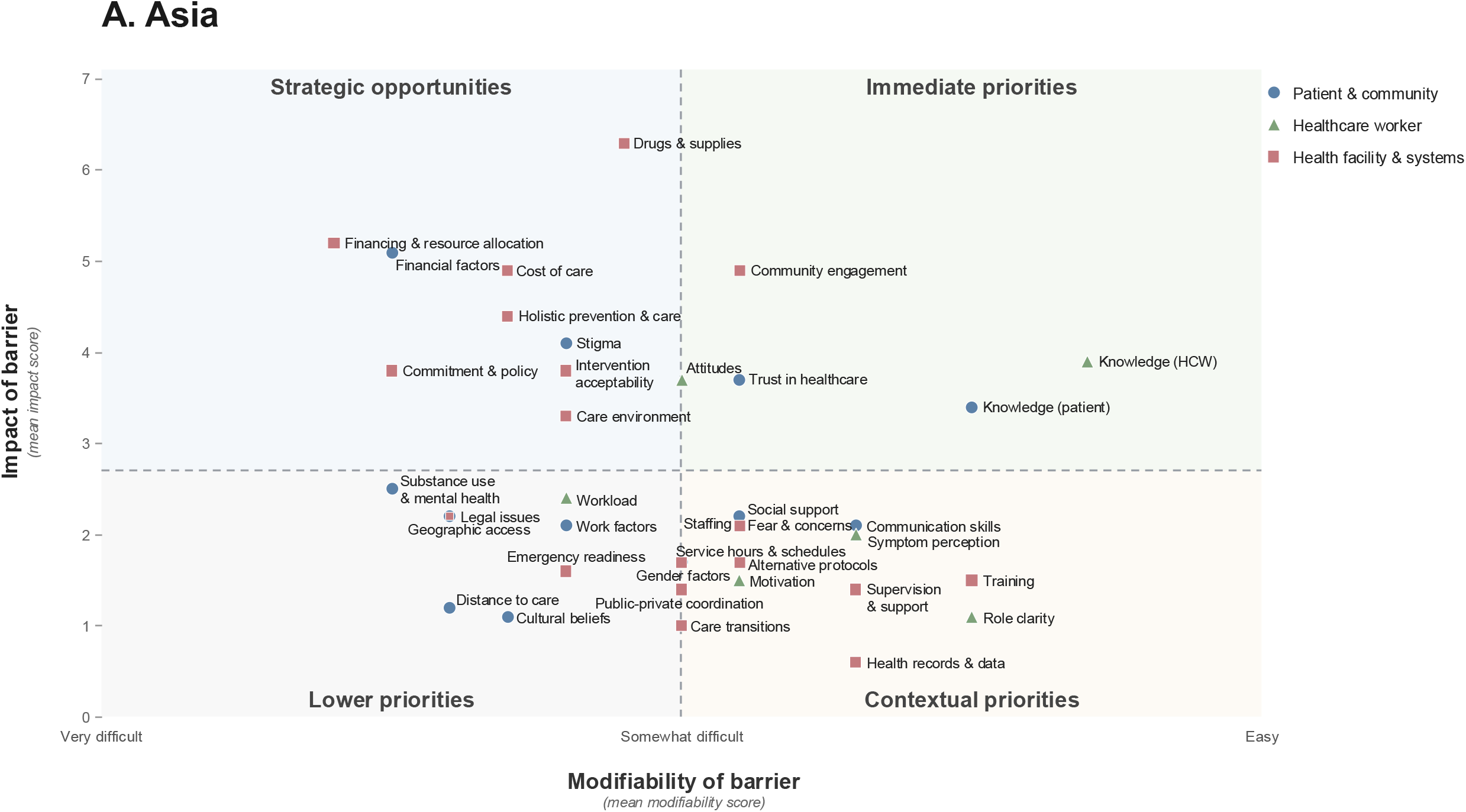
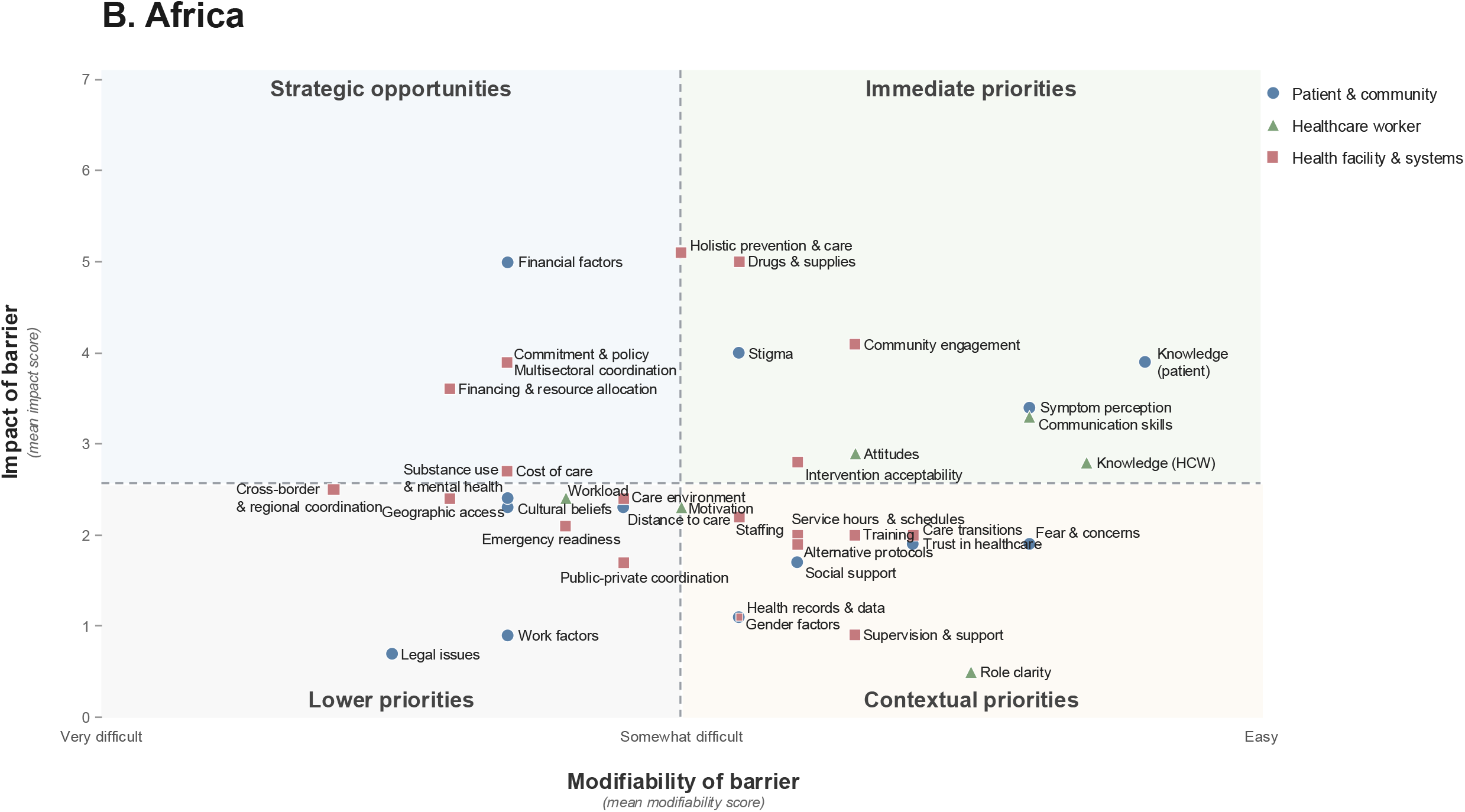
TB barrier priority matrices for (A) Asia (n=32) and (B) Africa (n=33), combining the stakeholder-perceived impact (harm) and modifiability. Impact was derived from the BWS exercise and modifiability from Likert-scale ratings; a barrier was considered a priority on impact if its mean impact score (MIS) exceeded the overall mean and on modifiability if its mean Likert score was ≥3. Barriers prioritized on both dimensions were conceptualized as immediate opportunities for implementation strategies (“immediate priorities”); those prioritized on impact but not modifiability as important but likely requiring longer-term or structural approaches (“strategic opportunities”); those prioritized on modifiability but not impact as addressable but lower-impact overall, warranting attention mainly in specific contexts where they are more harmful (“contextual priorities”); and those prioritized on neither as lower priority for implementation efforts (“lower priorities”).

## Discussion

This stakeholder-engaged process yielded a more complete and practice-informed understanding of the barriers that undermine demand, access, and retention along the TB care cascade in high-burden settings. By centering the voices of TB survivors, community advocates, and frontline HCWs (voices that rarely shape global TB priority-setting),^23,24^ we identified many barriers missing from prior reviews and developed a pragmatic framework for prioritizing barriers to address in resource-constrained settings. The resulting priorities reflect how TB care is experienced and delivered on the ground, rather than how barriers appear in the published literature, and provide targets for high-impact strategies to improve TB outcomes.

Participants across Asia and Africa converged on the barriers perceived as most harmful to good TB outcomes (7 of the top 10 shared across regions), despite substantial contextual differences. Challenges related to drug and supply reliability, financing and resource constraints, and the absence of holistic, person-centered care surfaced as dominant barriers. The workshops occurred against the backdrop of substantial disruptions to global TB funding in 2025,^25,26^ which may have influenced stakeholders’ perceptions. However, the cross-regional consistency of these barriers suggests that certain system-level constraints remain universally limiting regardless of regional structures or epidemiology. Stakeholder-driven refinement also highlighted barriers previously underrepresented in the literature, especially those related to coordination, governance, and community engagement. Notably, 7 out of 8 newly identified barriers were at the health system level, suggesting that the published literature adequately captures individual-level factors but underrepresents structural constraints highly salient to those delivering and receiving TB care.

Prioritization matrices provide a structured approach to differentiate between high-impact barriers that programs can address now, from those requiring longer-term investment. Four barriers emerged as consensus priorities for immediate action across both regions: patient/community knowledge, HCW knowledge, HCW attitudes, and lack of community engagement mechanisms. Because these span all three levels, they reinforce the need for coordinated, multilevel implementation strategies rather than isolated ones. Beyond these, regional variation emerged: Africa’s immediate priority quadrant additionally included systems-level drugs and supplies, holistic prevention and care, and intervention acceptability, patient/community-level stigma, and symptom perception, and HCW communication skills, while Asia’s included patient/community-level trust in healthcare. Such variation likely reflects differences in cultural norms, available resources, perceptions of government support, and prior exposure to successful care-improvement strategies. Nonetheless, the inclusion of both individual- and systems-level priorities across both regions demonstrates that communities and frontline HCWs are keenly aware of structural constraints, not solely of interpersonal interactions.

The barrier prioritization matrices should not be interpreted rigidly. While barriers in the immediate priority quadrant represent clear targets, several high-impact barriers in the strategic opportunities quadrant also warrant attention, despite being perceived as less modifiable — particularly in Asia, where 7 of the 10 highest-impact barriers fell into the strategic opportunities quadrant versus 4 in Africa. Patient financial factors, costs of care, financing and resource allocation, and TB commitment and policy fell into this quadrant in both regions; these are barriers where change is harder but the potential impact is substantial. Holistic prevention and care and intervention acceptability illustrate regional variation, perceived as changeable in Africa (immediate priorities) quadrant) but less modifiable in Asia (strategic opportunities). The structural and policy changes these barriers require, along with shifts in how interventions are developed and delivered, typically take longer than standard project cycles allow and are difficult to decouple from deeply entrenched practice,^27^ so sustained commitment is likely necessary to achieve meaningful improvements.^1,3^ Notably, even these barriers were rated as at least somewhat modifiable, suggesting stakeholders saw change as achievable. Programs must therefore weigh not only where change is easiest, but where it matters most.

TB programs have long emphasized individual-level behavior change strategies, such as health education, adherence support, and provider training, and stakeholders confirmed the importance of knowledge and attitudes. Most programs, however, already conduct TB sensitization and outreach to improve community knowledge and address symptom perception, and conduct training to address HCW knowledge and attitudes; this suggests the issue is not whether such activities are happening but how well they are operationalized. ^5^ Programs may need to reconceptualize how these strategies are delivered (e.g., who delivers them, in what format, how often, and with what content) rather than simply doing more of the same, to improve their reach, resonance, and impact.

This study should not be interpreted as defining the “true” priorities for any specific country or program. Rather, it provides a comprehensive, stakeholder-validated set of 39 barriers that can serve as a global reference for programs to systematically consider in their own settings, alongside a structured approach for prioritization based on local circumstances, resources, and policy windows. These findings serve as a foundation for TB-DASH,^28^ a global framework and tool to enhance TB demand, access, and care-seeking by modifying priority barriers by linking them to curated and highly promising evidence-based implementation strategies (i.e., “change strategies”).^29,30^ The tool aims to help programs move from identifying priority barriers to actionable, feasible, and context-specific strategies that can accelerate progress in high-burden settings.

This study has several strengths. Participants from 28 countries across the two highest-burden regions (Asia and Africa) contributed perspectives that reshaped the barrier landscape in ways the literature alone could not reveal. The in-person workshop format provided dedicated time and space for meaningful deliberation and refinement, and the community technical advisors and UCSF TRAC Community Advisory Board played key roles throughout, strengthening accessibility, trust, and engagement. The study also has limitations. We deliberately prioritized the voices of TB survivors, community advocates, and frontline HCWs, but policymakers and senior program managers, who were engaged later, may have different perceptions of barrier importance and modifiability. The sequential workshop design introduced asymmetry in the barrier sets evaluated across regions, but substantial overlap in top-ranked barriers suggests that this did not meaningfully bias regional comparisons. English-language requirements, while pragmatically necessary, may have hindered full engagement for some participants. BWS and modifiability assessments capture perceptions rather than objective effects or actual feasibility; these perceptions remain essential for designing realistic strategies but may not fully align with epidemiologic evidence or structural constraints. While the number of participants is modest, the depth of engagement, including two-day hybrid workshops with dedicated time for deliberation, refinement, and prioritization, yielded richer insights than larger, more superficial surveys could. Finally, while participants represented many diverse high-TB burden countries across Asia and Africa, findings may not generalize to Eastern Europe, Latin America, or the Western Pacific.

In conclusion, engaging TB survivors, community members, and frontline HCWs produced a more complete and actionable understanding of the barriers undermining TB prevention and care in Asia and Africa, demonstrating that centering these voices is not merely an ethical imperative but also essential for comprehensively identifying and prioritizing barriers that matter most. The priority barriers identified span patient/community, HCW, and health system levels, highlighting both immediate opportunities for impact, where change is perceived as highly feasible, and system-level constraints that require sustained investment but could offer substantial rewards. As TB programs navigate difficult decisions about resource allocation amid shifting global health funding, community-centered, stakeholder-driven barrier prioritization offers a principled approach to focusing implementation efforts where they are most needed and most likely to succeed.

## Data Availability

All data produced in the present work are contained in the manuscript

## Acknowledgements

We thank UC TRAC TB Community Advisory Board members and our stakeholder participants who made crucial contributions to this work by sharing their expertise and lived experience.

## Author contributions

ADK and PS conceived and designed the study. KS, SRC, CFH conducted the scoping reviews that generated the initial barrier set. ADK, KS, PS, NK, MN, and KA designed and facilitated the regional stakeholder workshops. KS, NK, MN, KA, TS, EL, and PS led community engagement, participant curation, and refinement of barrier language for accessibility. ADK, KS, PS designed the BWS and modifiability exercises, while ADK conducted the analyses. ADK, KS, and PS drafted the initial manuscript. All authors contributed to data interpretation, critically reviewed and revised the manuscript, and approved the final version.

## Box 1.

**Stakeholder workshop design and procedures**

**Workshop preparation**

Before the regional workshops, we refined barrier language to ensure accessibility to diverse stakeholders, including those without technical or medical backgrounds. First, an artificial intelligence (AI) model (Claude, Anthropic) provided a plain-language review (at an 8th-grade reading level) to identify overly technical terms or concepts and suggest accessible alternatives; all AI-suggested revisions were reviewed by the research team before adoption. The University of California, San Francisco Tuberculosis Research Advancement Center (UCSF TRAC) TB Community Advisory Board, largely comprised of TB survivors and community advocates from high-burden countries, then reviewed the barriers for resonance and appropriateness through a virtual-guided exercise on Zoom.

**Participant recruitment and selection**

We deliberately prioritized participation from groups typically underrepresented in TB policy discussions: TB survivors, community advocates, and frontline HCWs. This intentional focus reflected our aim to capture diverse perspectives that may be missing from the published literature, which is often shaped by the viewpoints of researchers. Participants were recruited purposively through TB advocacy networks, community-based organizations, national TB programs, and the research team’s professional networks. Eligibility criteria included direct experience with TB (as a survivor, caregiver, community advocate, or healthcare provider) or expertise in TB program implementation; residence or primary work in a high TB burden country in the relevant region; and ability to participate in English-language discussions. We carefully curated the list of participant invitations to ensure geographic representation across multiple countries within each region, balance across stakeholder types (TB survivors, community advocates, frontline HCWs, program implementers), relevant expertise distributed across the three barrier levels, and a mix of perspectives from different healthcare settings, including public and private sectors and urban and rural areas. Enrollment was capped at approximately 30 in-person participants per workshop to enable meaningful participation and discussion. The UCSF TRAC TB CAB emphasized the importance of including frontline HCWs and community members from rural areas, while final participant selection was guided by the research team and embedded community technical advisors (TB survivors and experienced advocates, residing in high-burden countries). To ensure meaningful participation and reduce financial barriers to engagement, we covered all travel, accommodation, and meal costs for in-person participants. A virtual participation option was offered for individuals unable to travel.

**Workshop structure**

Day 1 for each workshop comprised three parts: presentation of the barriers, participatory refinement and expansion, and prioritization. First, the study team community technical advisors presented the barriers one by one, grouped by theme and level, with opportunities for clarification and questions throughout. To accompany this process, participants received a barrier reference guide describing each of the 31 literature-derived barriers, including its definition in lay language, the TB care cascade step(s) it most directly affects, and its influence on TB prevention and care outcomes (**Supplementary File 1**). To refine and expand the barrier list, in-person and online participants were divided into three groups corresponding to the barrier levels (patient/community, HCW, health system), each assigned to the level most aligned with their lived or professional experience. During a 30-minute session, groups reviewed barriers assigned to their level and documented proposed refinements, additions, or consolidations on large poster paper. This was followed by a 30-minute crosswalk in which each group rotated to review, discuss, and further refine the barriers of the other two levels. Facilitators compiled proposed additions and refinements, and each group was then given dedicated time to report back their high-level findings to the larger group. New barriers meeting group consensus and deemed distinct from the literature-derived set were added to the final barrier set for further prioritization.

**Barrier assessment and real-time results sharing**

Following the refinement process, participants completed a Best-Worst Scaling exercise and modifiability assessment of the revised barrier set. A short presentation explained both assessment formats and emphasized the importance of honest feedback. Participants had one hour to complete the assessments on their personal devices or on computers provided by the research team. Technical support was available throughout (for both in-person and online participants), allowing for the clarification of any questions about barrier definitions. BWS and modifiability data were analyzed immediately following survey completion, and preliminary findings were presented to workshop participants before the end of Day 1. Real-time results sharing enabled stakeholder verification that the findings aligned with their collective understanding, facilitated discussion and co-interpretation of the results with the community that generated the data, and increased transparency and ownership of the findings.

