## Supplementary Tables for "Identifying and Prioritizing Barriers to TB Prevention and Care in High-Burden Countries: A Community-Engaged Approach Using Best-Worst Scaling"

**Supplementary Table 1. Number of participants by country and WHO TB high-burden country designation (2021-2025)**

| **Country** | **Total workshop participants**  **(n=81)** | **Participants completing the survey-based barrier assessment**  **(n=65)** | **WHO TB high-burden country designation** |
| --- | --- | --- | --- |
| **Asia Workshop** | | | |
| **Bangladesh** | 2 | 2 | TB; MDR/RR-TB |
| **Cambodia** | 1 | 1 | None |
| **India** | 13 | 9 | TB; TB/HIV; MDR/RR-TB |
| **Indonesia** | 2 | 1 | TB; TB/HIV; MDR/RR-TB |
| **Myanmar** | 3 | 2 | TB; TB/HIV; MDR/RR-TB |
| **Nepal** | 3 | 3 | MDR/RR-TB |
| **Pakistan** | 1 | 0 | TB; MDR/RR-TB |
| **Philippines** | 5 | 3 | TB; TB/HIV; MDR/RR-TB |
| **Sri Lanka** | 1 | 1 | None |
| **Thailand** | 3 | 2 | TB; TB/HIV |
| **Vietnam** | 9 | 8 | TB; MDR/RR-TB |
| **Africa Workshop** | | | |
| **Botswana** | 2 | 1 | TB/HIV |
| **Cote D'Ivoire** | 1 | 1 | None |
| **Eswatini** | 1 | 1 | TB/HIV |
| **Ethiopia** | 2 | 2 | TB; TB/HIV |
| **Ghana** | 1 | 1 | None |
| **Kenya** | 4 | 3 | TB; TB/HIV |
| **Lesotho** | 1 | 1 | TB; TB/HIV |
| **Liberia** | 1 | 1 | TB; TB/HIV |
| **Malawi** | 4 | 4 | TB/HIV |
| **Mozambique** | 1 | 1 | TB; TB/HIV; MDR/RR-TB |
| **Nigeria** | 3 | 2 | TB; TB/HIV; MDR/RR-TB |
| **Rwanda** | 1 | 1 | None |
| **South Africa** | 6 | 6 | TB; TB/HIV; MDR/RR-TB |
| **Tanzania** | 1 | 0 | TB; TB/HIV |
| **Uganda** | 6 | 5 | TB; TB/HIV |
| **Zambia** | 1 | 1 | TB; TB/HIV; MDR/RR-TB |
| **Zimbabwe** | 2 | 2 | TB/HIV; MDR/RR-TB |

**Supplementary Table 2. Characteristics of workshop participants who completed the survey-based barrier assessment**

|  | **Overall (n=65)** | **Asia Workshop (n=32)** | **Africa Workshop (n=33)** |
| --- | --- | --- | --- |
| **Countries represented** | 26 | 10 | 16 |
| **Age group** |  |  |  |
| <34 years | 12 (18) | 4 (13) | 8 (24) |
| 35-54 | 42 (65) | 20 (63) | 22 (67) |
| 55+ | 11 (17) | 8 (25) | 3 (9) |
| **Sex^^^** |  |  |  |
| Female | 31 (49) | 12 (40) | 19 (58) |
| Male | 32 (51) | 18 (60) | 14 (42) |
| **TB survivor*** |  |  |  |
| Yes | 11 (17) | 6 (19) | 5 (16) |
| No | 53 (83) | 26 (81) | 27 (84) |
| **Primary role** |  |  |  |
| Community Representative | 16 (25) | 6 (19) | 10 (30) |
| Frontline healthcare worker | 23 (35) | 15 (47) | 8 (24) |
| Policy/Decision Maker | 13 (20) | 6 (19) | 7 (21) |
| Researcher | 10 (15) | 5 (16) | 5 (15) |
| Other | 3 (5) | 0 | 3 (9) |
| **Years of TB work experience^#^** |  |  |  |
| 0-3 | 9 (14) | 5 (16) | 4 (12) |
| 4-10 | 19 (29) | 9 (28) | 10 (30) |
| 10-19 | 23 (35) | 9 (28) | 14 (42) |
| 20+ | 14 (22) | 9 (28) | 5 (15) |
| **Primary setting*** |  |  |  |
| Urban | 23 (36) | 11 (35) | 12 (36) |
| Rural | 4 (6) | 1 (3) | 3 (9) |
| Both | 37 (58) | 19 (61) | 18 (55) |
| **Involved in national TB decision making^^^** |  |  |  |
| Yes | 27 (43) | 11 (35) | 16 (50) |
| No | 36 (57) | 20 (65) | 16 (50) |

^#^Includes both paid and volunteer work. ^^^2 people preferred not to report; *1 person preferred not to report

**Supplementary Table 3. TB barrier impact and modifiability scores by region**

| **Barrier** | **Level** | **Asia (n=32)** | | | | **Africa (n=33)** | | | |
| --- | --- | --- | --- | --- | --- | --- | --- | --- | --- |
|  |  | **Mean Impact Score** | **Impact Rank**  **(of 37)** | **Mean Modifiability Score** | **Priority category** | **Mean Impact Score** | **Impact Rank**  **(of 39)** | **Mean Modifiability Score** | **Priority category** |
| Cultural beliefs | Patient | 1.1 (0.6-1.7) | 35 | 2.7 (2.4-2.9) | Lower | 2.3 (1.7-2.8) | 23 | 2.7 (2.4-3.0) | Lower |
| Distance to care | Patient | 1.2 (0.7-1.7) | 33 | 2.6 (2.2-2.9) | Lower | 2.3 (1.6-3.0) | 22 | 2.9 (2.6-3.2) | Lower |
| Fear & concerns | Patient | 2.2 (1.6-2.8) | 20 | 3.1 (2.9-3.4) | Contextual | 1.9 (1.2-2.5) | 31 | 3.6 (3.4-3.8) | Contextual |
| Financial factors | Patient | 5.1 (4.2-5.9) | 3 | 2.5 (2.3-2.7) | Strategic | 5.0 (4.3-5.7) | 3 | 2.7 (2.4-3.0) | Strategic |
| Gender factors | Patient | 1.7 (1.1-2.4) | 25 | 3.0 (2.8-3.2) | Contextual | 1.1 (0.6-1.7) | 34 | 3.1 (2.8-3.4) | Contextual |
| Knowledge | Patient | 3.4 (2.6-4.1) | 13 | 3.5 (3.3-3.8) | Immediate | 3.9 (3.1-4.7) | 7 | 3.8 (3.7-4.0) | Immediate |
| Legal issues | Patient | 2.2 (1.3-3.1) | 19 | 2.6 (2.3-2.9) | Lower | 0.7 (0.3-1.1) | 38 | 2.5 (2.2-2.8) | Lower |
| Social support | Patient | 2.2 (1.4-3.0) | 18 | 3.1 (2.9-3.3) | Contextual | 1.7 (1.0-2.4) | 33 | 3.2 (3.0-3.4) | Contextual |
| Stigma | Patient | 4.1 (3.2-5.0) | 7 | 2.8 (2.5-3.0) | Strategic | 4.0 (3.3-4.8) | 5 | 3.1 (2.8-3.4) | Immediate |
| Substance use & mental health | Patient | 2.5 (1.0-3.2) | 15 | 2.5 (2.2-2.7) | Lower | 2.4 (1.5-3.2) | 19 | 2.7 (2.5-2.9) | Lower |
| Symptom perception | Patient | 2.1 (1.4-2.8) | 22 | 3.3 (3.1-3.5) | Contextual | 3.4 (2.6-4.3) | 10 | 3.6 (3.4-3.8) | Immediate |
| Trust in healthcare | Patient | 3.7 (2.9-4.6) | 11 | 3.1 (2.9-3.3) | Immediate | 1.9 (1.4-2.3) | 30 | 3.4 (3.1-3.6) | Contextual |
| Work factors | Patient | 2.1 (1.4-2.8) | 23 | 2.8 (2.5-3.1) | Lower | 0.9 (0.5-1.2) | 37 | 2.7 (2.4-3.0) | Lower |
| Attitudes | HCW | 3.7 (2.8-4.5) | 12 | 3.0 (2.8-3.2) | Immediate | 2.9 (2.1-3.6) | 12 | 3.3 (3.1-3.6) | Immediate |
| Communication skills | HCW | 2.0 (1.3-2.6) | 24 | 3.3 (3.1-3.6) | Contextual | 3.3 (2.6-4.1) | 11 | 3.6 (3.4-3.8) | Immediate |
| Knowledge | HCW | 3.9 (3.2-4.6) | 8 | 3.7 (3.5-3.9) | Immediate | 2.8 (2.1-3.5) | 14 | 3.7 (3.5-3.9) | Immediate |
| Motivation | HCW | 1.5 (1.0-2.1) | 29 | 3.1 (2.8-3.3) | Contextual | 2.3 (1.6-3.1) | 21 | 3.0 (2.7-3.2) | Contextual |
| Role clarity | HCW | 1.1 (0.6-1.6) | 34 | 3.5 (3.2-3.7) | Contextual | 0.5 (0.3-0.7) | 39 | 3.5 (3.3-3.7) | Contextual |
| Workload | HCW | 2.4 (1.6-3.2) | 16 | 2.8 (2.5-3.0) | Lower | 2.4 (1.8-3.0) | 20 | 2.8 (2.6-3.0) | Lower |
| Alternative protocols | Systems | 1.7 (1.2-2.2) | 26 | 3.1 (2.8-3.3) | Contextual | 2.0 (1.5-2.4) | 27 | 3.2 (3.0-3.5) | Contextual |
| Care environment | Systems | 3.3 (2.5-4.0) | 14 | 2.8 (2.5-3.0) | Strategic | 2.4 (1.8-3.1) | 18 | 2.9 (2.7-3.2) | Lower |
| Care transitions | Systems | 1.0 (0.6-1.4) | 36 | 3.0 (2.7-3.3) | Contextual | 2.0 (1.4-2.7) | 26 | 3.4 (3.2-3.7) | Contextual |
| Commitment & policy | Systems | 3.8 (2.8-4.8) | 10 | 2.5 (2.2-2.7) | Strategic | 3.9 (3.1-4.7) | 6 | 2.7 (2.4-3.0) | Strategic |
| Community engagement | Systems | 4.9 (4.1-5.8) | 4 | 3.1 (2.8-3.3) | Immediate | 4.1 (3.3-5.0) | 4 | 3.3 (3.1-3.5) | Immediate |
| Cost of care | Systems | 4.9 (4.1-5.8) | 5 | 2.7 (2.5-3.0) | Strategic | 2.7 (1.9-3.5) | 15 | 2.7 (2.4-3.0) | Strategic |
| Cross-border & regional coordination | Systems | - | - | - | - | 2.5 (1.8-3.3) | 16 | 2.4 (2.1-2.6) | Lower |
| Drugs & supplies | Systems | 6.3 (5.6-7.0) | 1 | 2.9 (2.6-3.2) | Strategic | 5.0 (4.2-5.9) | 2 | 3.1 (2.9-3.4) | Immediate |
| Emergency readiness | Systems | 1.6 (1.0-2.2) | 28 | 2.8 (2.5-3.0) | Lower | 2.1 (1.3-2.8) | 25 | 2.8 (2.6-3.0) | Lower |
| Financing & resource allocation | Systems | 5.2 (4.5-5.9) | 2 | 2.4 (2.2-2.6) | Strategic | 3.6 (3.0-4.2) | 9 | 2.6 (2.4-2.8) | Strategic |
| Geographic access | Systems | 2.2 (1.5-2.9) | 17 | 2.6 (2.3-3.0) | Lower | 2.4 (1.7-3.2) | 17 | 2.6 (2.3-2.9) | Lower |
| Health records & data | Systems | 0.6 (0.4-0.8) | 37 | 3.3 (2.0-2.5) | Contextual | 1.1 (0.6-1.5) | 35 | 3.1 (2.9-3.4) | Contextual |
| Holistic prevention & care | Systems | 4.4 (3.6-5.1) | 6 | 2.7 (2.5-2.9) | Strategic | 5.1 (4.2-6.0) | 1 | 3.0 (2.7-3.3) | Immediate |
| Intervention acceptability | Systems | 3.8 (3.1-4.6) | 9 | 2.8 (2.6-3.0) | Strategic | 2.8 (2.1-3.5) | 13 | 3.2 (2.9-3.5) | Immediate |
| Multisectoral coordination | Systems | - | - | - | - | 3.9 (3.0-4.7) | 8 | 2.7 (2.4-3.0) | Strategic |
| Public-private coordination | Systems | 1.4 (0.9-2.0) | 31 | 3.0 (2.7-3.3) | Contextual | 1.7 (1.0-2.5) | 32 | 2.9 (2.6-3.3) | Lower |
| Service hours & schedules | Systems | 1.7 (1.0-2.3) | 27 | 3.0 (2.7-3.3) | Contextual | 1.9 (1.3-2.5) | 29 | 3.2 (3.0-3.5) | Contextual |
| Staffing | Systems | 2.1 (1.4-2.8) | 21 | 3.1 (2.8-3.3) | Contextual | 2.2 (1.5-3.0) | 24 | 3.1 (2.8-3.3) | Contextual |
| Training | Systems | 1.5 (0.9-2.0) | 30 | 3.5 (3.3-3.8) | Contextual | 2.0 (1.5-2.4) | 28 | 3.3 (3.0-3.5) | Contextual |
| Supervision & support | Systems | 1.4 (0.8-1.9) | 32 | 3.3 (3.0-3.5) | Contextual | 0.9 (0.3-1.5) | 36 | 3.3 (3.1-3.6) | Contextual |

Mean impact scores represent BWS-derived mean preference weights and sum to 100 across all barriers. Mean modifiability scores range from 1 (“not modifiable”) to 4 (easy to

modify). Barriers exceeding the workshop-specific overall mean (Asia: 2.70; Africa: 2.56) were classified as a priority based on impact.
